# Syndemic violence victimization, alcohol and drug use, and HIV transmission risk behavior among transgender women in India: A cross-sectional, population-based study

**DOI:** 10.1101/2022.04.07.22273584

**Authors:** Venkatesan Chakrapani, P.V.M. Lakshmi, Peter A. Newman, Jasvir Kaur, Alexander C. Tsai, P.P. Vijin, Bhawani Singh, Pradeep Kumar, Shobini Rajan, Rajesh Kumar

## Abstract

**Introduction:** Transgender women are disproportionately burdened by HIV. Co-occurring epidemics of adverse psychosocial exposures accelerate HIV sexual risk, including among transgender women; however, studies using additive models fail to examine synergies among psychosocial conditions that define a syndemic. We examined the impact of synergistic interactions among 4 psychosocial exposures on condomless anal sex (CAS) among a national probability sample of transgender women in India.

**Methods:** A probability-based sample of 4,607 HIV-negative transgender women completed the Indian Integrated Bio-behavioral Surveillance survey, 2014−2015. We used linear probability regression and logistic regression to assess 2-, 3-, and 4-way interactions among 4 exposures (physical and sexual violence, drug and alcohol use) on CAS.

**Results:** Overall, 27.3% reported physical and 22.3% sexual violence victimization (39.2% either physical or sexual violence), one-third (33.9%) reported frequent alcohol use and 11.5% illicit drug use. Physical violence was associated with twofold higher odds of CAS in the main effects model. Significant two- and three-way interactions were identified on both multiplicative and additive scales between physical violence and drug use; physical and sexual violence; physical violence, sexual violence, and alcohol use; and physical violence, alcohol and drug use.

**Conclusions:** Physical and sexual violence victimization, and alcohol and drug use are highly prevalent and synergistically interact to increase CAS among transgender women in India. Targeted and integrated initiatives to improve assessment of psychosocial comorbidities, to combat transphobic violence, and to provide tailored, trauma-informed alcohol and substance use treatment services may reduce HIV risk among transgender women.

## Introduction

Transgender women globally bear a disproportionate HIV burden [1] secondary to rampant stigma and discrimination in various domains of their lives [2]. India’s National AIDS Control Organization (NACO) designates transgender women a ‘high risk group’ or ‘key population’, with official HIV prevalence estimates ranging from 9.5% (in 2015 [3]) to 3.1% (in 2017 [4])—from 14 to over 40 times higher than the general population (0.22%) [5]. Until 2006, NACO-supported HIV preventive interventions subsumed transgender women under “men who have sex with men”; but they are now reached through 41 interventions exclusively for transgender women and 152 ‘core-composite’ targeted HIV interventions for transgender women with other at-risk populations [5]. These targeted interventions focus on HIV and STI education, condom promotion, free condom distribution, and HIV testing referrals. Transgender women in India have demonstrated high (89%) awareness of the usefulness of condoms in preventing HIV infection; nevertheless, consistent condom use remains low at 45% to 54% [4,6].

Syndemic theory has been applied to explain the clustering and concentration of diseases such as HIV in vulnerable populations [7], including transgender women [1,8]. In their classic paper [9], Stall et al. highlighted the public health impacts of co-occurring psychosocial problems among gay men in the U.S., advancing a life-course theory of syndemic production rooted in the cultural marginalization of a population that is heavily stigmatized [10]. In the Indian context, harmful social conditions such as societal stigma faced by transgender women may lead to clustering of psychosocial health problems, such as violence victimization, problematic alcohol use, depression, and HIV risk [11,12]; these may mutually reinforce one another and synergistically amplify disease burden.

Transgender women in India have a high prevalence of psychosocial health problems, including lifetime physical or sexual violence victimization (84.0%), alcohol use (37.3%) and depression (35.3%), all of which have demonstrated significant additive effects on HIV risk [13]. One study reported 18% past-year prevalence of sexual violence victimization, with higher risk for HIV among those who experienced sexual violence [14].

Exploring the presence of synergy among psychosocial health conditions requires the use of appropriate analytical strategies. A recommended analytical technique for testing synergy is by checking for statistical interaction (on the additive and/or multiplicative scale) between co-occurring conditions [15,16]. Most analyses in western literature that purportedly explored synergy used the number of psychosocial conditions or exposures as a cumulative count to predict HIV risk behavior, which does not address the interaction concept embedded in syndemic theory [15]. Two papers on syndemics among transgender women in India each used analytical strategies that did not involve testing for interactions between psychosocial conditions. One study used the number of psychosocial syndemic conditions as a cumulative count to predict HIV risk behavior [13]. Another study used latent class analysis to identify different ‘syndemic classes’ (combinations of depression, alcohol use, and/or violence victimization), which were associated with higher HIV risk compared to no syndemic class [6].

To address research gaps in the application of syndemic theory and support the design of tailored, evidence informed interventions, we assessed potential synergistic interactions between sexual and physical violence, alcohol use, and drug use on condomless anal sex (CAS) among transgender women.

## Materials and Methods

### Study design and population

Data for these analyses were drawn from a population-based cross-sectional survey among transgender women recruited under India’s National AIDS Control Organization (NACO) Integrated Bio-Behavioral Surveillance (IBBS) study, last conducted from 2014−2015. The IBBS aimed to generate evidence on risk behaviors among groups deemed high-risk (e.g., transgender women, female sex workers, men who have sex with men) to support planning and prioritization of HIV program efforts. Eligibility criteria were as follows: persons aged ≥15 years, whose self-identity does not conform unambiguously to conventional notions of male or female gender roles but combines or moves between them.

The IBBS study was approved by the ethics committees of NACO and participating Indian Council of Medical Research regional institutes. All participants provided written informed consent; for those 15-17 years-old, consent was also obtained from their parent/guardian. Monetary compensation of INR 200 (∼3 USD) was provided.

This paper presents findings from secondary analysis of NACO population-based study data to examine the burden of adverse psychosocial exposures, co-occurrence of these exposures, and their potential independent and synergistic effects on CAS among transgender women.

### Sampling and recruitment

The IBBS sample size was calculated with the primary objective to measure changes in consistent condom use with sexual partners and changes in HIV prevalence. The primary unit of survey was a ‘domain’: a geographical unit of either a single district or a group of four socio-culturally similar districts where a single district had an insufficient sample size. Sampling including both conventional cluster sampling (CCS)—in sites (e.g., homes, brothels) where transgender women could be found at any time of the day, and time-location cluster sampling (TLCS)—in sites such as parks and beaches where transgender women could be found on particular days and times to meet or socialize with other transgender people or potential male partners. The estimated sample size of transgender women for each domain was >700. However, considering feasibility based on sample availability in different domains, a target sample size of 400 per domain was chosen. In domains with sample sizes <400, a ‘take all’ approach was used. Overall, the target sample size was 5,588 transgender women in 15 domains across 11 states. A valid sample of 4,966 transgender women was achieved, with an 81% response rate. NACO’s online report [17] provides further details on sampling and recruitment.

### Measures

Information on participants’ sociodemographic characteristics included age (in years), years of education, marital status, engagement in sex work (past year) and self-identified gender-affirmative surgical status (*ackwa*: pre-or non-operative, or *nirvan:* post-operative). Table 1 describes how the outcome, primary exposures, and other relevant covariates used in the analysis were measured—survey items, response range and scoring, and creation of binary and continuous measures. The binary outcome measure was CAS with any type (regular, paying, paid, and casual) of male sexual partner in the past month. Binary adverse psychosocial exposures included physical violence [PV], sexual violence [SV], and drug use [D] (non-injection or injection drugs) in the past year, and frequent alcohol use [A] (>3 days) in the past week (Table 1). Other covariates included engagement in sex work, forced sex experience during adolescence (<18 years), HIV transmission knowledge (score), HIV risk perception (high vs. low), social support (score), and HIV program exposure (score).

**Table 1.** Summary of Key Measures: Outcome, Primary Exposures, and other Relevant Covariates.

| Measures | Survey items |  | Range of responses, coding, and scores | Types of variables |
| --- | --- | --- | --- | --- |
|  | Number of items | Questions asked |  |  |
| **Outcome** |  |  |  |  |
| *Condomless anal sex with male partners* (past month) | 4 | In the last one month, how often have you used condoms when you had anal sex with your ...?<br>a. regular non-paying male partner<br>b. paying male partner<br>c. paid male partner<br>d. casual non-paying male partners | 0 for 'every time' and 1 for all other options (most of the time, sometimes, never)<br>'Condomless anal sex' was created by combining the condom use (=1) responses from these four types of partners. | Binary |
| **Primary exposures** |  |  |  |  |
| *Physical violence* (past year) | 2 | In the last 12 months, how many times would you say someone has beaten (hurt, hit, slapped, pushed, kicked, punched, choked or burned) you?<br>In the last 12 months, who was the person (or people) who have beaten you? | 0 for 'never' and 1 for other responses (Once, 2-5 times, 6-10 times, >10 times).<br>Those who responded as "Do not remember" but reported the person who caused physical violence were also categorized as having experienced physical violence. | Binary |
| *Sexual violence* (past year) | 1 | In the last 12 months, were you physically forced to have sexual intercourse with someone even though you didn't want to? | Yes=1, No=0 | Binary |
| *Violence victimization* (past year) | - | Recoding of the final coding of physical and sexual violence variables. | 'Violence victimization' was created by considering the codes of physical violence (from never=0, Once=1, 2-5 times=2, 6-10 times=3, >10 times=4) and sexual violence (Yes=1, No=0) as scores, and combining these scores.<br>Score range: 0-5 | Continuous |
| *Alcohol use* (past week) | 1 | How many days did you consume alcohol in the last one week? | Responses dichotomized at the upper quartile of consumption, $\leq 3$ vs. $>3$ days/week.<br>Score range: 0-7<br>(Number of days of alcohol use in the past week) | Binary<br>Continuous |
| *Drug use* (past year) | 2 | Have you consumed drugs such as Ganja, Heroine for pleasure in the last 12 months?<br>Have you injected drugs for nonmedical reasons in the last 12 months? | Yes=1, No=0<br>Consumption of any drug (injection or not) was considered as 'drug use'.<br>Score range: 0-3 | Binary<br>Continuous |
| **Relevant covariates** |  |  |  |  |
| *Engagement in sex work (past year)* | 1 | Have you received cash or gifts from men in exchange for sex in the last 12 months? | Yes=1, No=0 | Binary |
| *Knowledge of HIV transmission risk* | 5 | Can a person get HIV/AIDS?<br>a. by having unprotected sex with an infected person<br>b. by sharing infected needles<br>c. by infected blood transfusion<br>d. through mosquito bites<br>Can HIV be transmitted from an HIV infected mother to<br>e. her unborn baby during pregnancy/delivery or to the new born child through breastfeeding? | Yes=1, No=0<br>For the analysis, each correct response was coded as 1, otherwise 'zero'.<br>Score range: 0 to 5 | Continuous |
| *HIV risk perception* | 1 | To what extent do you feel yourself at risk to being infected with HIV/AIDS? | High=1, Moderate=2, Low=3, No risk=4<br>Responses were dichotomized by combining response 1 and 2 as 'High risk', 3 and 4 as 'Low risk' | Binary |
| *Forced sex experience during adolescence* | 2 | How old were you when you had your first sex with a male?<br>Were you forced to have sex during the first sexual encounter with a male? | Participants who reported first sex with a male when they were <18 years (age of consent for heterosexual intercourse in India), and of them who reported forced sex during the first sexual encounter with a male, were categorised as having experienced forced sex during adolescence. | Binary |
| *HIV program exposure* | 8 | Have you received following services from any NGO/program/individual/group during the last 12 months?<br>a. Received information on STI/HIV/AIDS<br>b. Received condoms<br>c. Received lubricants<br>d. Seen a demonstration on correct condom use<br>e. Received check-up and counselling for STIs<br>f. Received free medicine for STIs<br>g. Visited drop in center<br>h. Referred to other services (STI clinic, HIV testing, etc.) | Yes=1, No=0<br>Score range: 0 to 8 | Continuous |
| *Social support score* | 3 | Have you received following services from any NGO/program/individual/group during the last 12 months?<br>a. Received help and support when faced with physical or sexual violence<br>b. Received help and support when faced with trouble from police<br>Are you a member of ...?<br>c. a self-help group or transgender women collective | Yes=1, No=0<br>Score range: 0 to 3 | Continuous |

### Analysis

The present analysis was restricted to HIV-negative transgender women (n=4,607). HIV-positive participants (n=359; 7.2%) were excluded given the objective of identifying correlates of HIV risk behavior to inform primary prevention initiatives. All analyses were conducted in Stata (version 16; College Station, Texas, USA). Descriptive statistics were used to summarize participant characteristics. Independent associations between adverse psychosocial exposures (binary) and CAS (binary) were estimated using logistic regression. The extent to which the adverse psychosocial exposures synergistically interacted with each other in their associations with CAS was examined by assessing interactions on the additive scale using linear probability regression models, and on the multiplicative scale using logistic regression models [15,18]. All possible two-way (e.g., physical violence x drug use), three-way (e.g., sexual violence x drug use x alcohol use), and four-way interaction terms (e.g., physical violence x sexual violence x drug use x alcohol use) were included in the regression models. All regression models were adjusted for age (in years), education, marital status (single vs. heterosexually married), gender-affirmative surgical status (ackwa vs. nirvan), engagement in sex work, forced sex experience during adolescence, knowledge of HIV transmission, HIV risk perception, social support, and exposure to HIV interventions. To account for the complex survey design [19] involving cluster sampling, ‘svyset’ and ‘svy’ Stata commands were used before estimating regression models [20].

On the additive scale in linear probability regression models, synergy is interpreted as statistically significant relative excess risk due to interaction (RERI) greater than zero; on the multiplicative scale in logistic regression models, synergy is interpreted as statistically significant coefficients on the product terms [18,21]. Multiplicative interaction estimated on the odds scale using logistic regression was re-estimated as ‘semi-elasticities’ on the probability scale using the Stata ‘margins’ command. The estimated semi-elasticities can be interpreted as the proportional change in the probability of CAS that is associated with a one unit change in the covariate or with the interaction. For example, a semi-elasticity of .40 is interpreted as a 40% relative change in the expected probability of CAS associated with a one unit change in the covariate (e.g., age in years) or with the joint effect of two (PV x D) or more (PV x SV x D) exposures, above and beyond their independent associations with the outcome.

### Sensitivity analysis for the model of synergistically interacting epidemics

To confirm the robustness of our findings, as part of the sensitivity analysis, we specified: 1) drug use (score range, 0–3) and alcohol use (range, 0–7) as continuous exposures; 2) physical and sexual violence (score range, 0–5) as a single continuous exposure, along with CAS (range, 0–4) as a continuous outcome in both sensitivity analyses.

### Exploratory analyses to check for rival theoretical models

In addition to synergistically interacting epidemics, Tsai [22] described two other models of co-occurring epidemics—the serially causal epidemics model and mutually causal epidemics model [23]—that have been used, variably, in the literature on syndemics. Although our analysis was focused specifically on testing a model of synergistically interacting epidemics, we also tested these other two models as they may suggest alternate or complementary mechanisms of risk. To test the model of serially causal epidemics, we conceptualized alcohol (score range, 0–7) and drug use (range, 0–3) as potential mediators of the association between sexual and physical violence victimization and CAS (range, 0–4). Similarly, we tested the model of mutually causal epidemics, in which we hypothesized that violence victimization (range, 0–5) and drug use (range, 0–3) were mutually reinforcing of one another, given that both were measured in the same time-period (past year). For assessing both of these models, we conducted path analysis (sem command) in Stata-16.

## Results

### Participant characteristics

Table 2 describes participant characteristics. Participants’ mean age was 28.5 years (SD 7.4) and mean number of years of education was 8.4 (SD 4.0). A majority (86.1%) were currently single. Nearly one-third (31.1%) reported sex work as their main occupation, while over half (56.8%) reported engaging in sex work during the past month. About one-fifth (18.3%) reported having experienced forced sex during adolescence (<18 years of age). More than half (57.2%) self-identified as ackwa, 42.7% as nirvan. About three-fifths (61.6%) perceived themselves at high risk for HIV infection. Reported CAS with different types of male partners in the past month ranged from 35.0% to 46.0%; CAS with any type of male partner was 45.1%.

**Table 2.** Characteristics of HIV-negative Transgender Women who Participated in the Integrated Bio-Behavioral Surveillance (IBBS) study, 2014-15 (N = 4,607)

| Characteristics | Mean (SD) |
| --- | --- |
| Age (in years) | 28.5 (7.4) |
| Education (years) | 8.4 (4.0) |
| HIV transmission knowledge score | 4.3 (1.2) |
| HIV program exposure score | 4.7 (2.8) |
| Social support score | 1.7 (1.0) |
|  | **N (%)** |
| Engagement in sex work (past month) | 2617 (56.8) |
| Gender-affirmative surgical status |  |
| Ackwa (pre-operative/non-operative) | 2636 (57.2) |
| Nirvan (post-operative) | 1969 (42.7) |
| Current marital status |  |
| Single (never married/widowed/divorced/separated) | 3940 (86.1) |
| Married | 637 (13.9) |
| HIV risk perception<sup>a</sup> |  |
| Low | 1594 (34.6) |
| High | 2839 (61.6) |
| Forced sex experience during adolescence | 837 (18.2) |
| Condomless anal sex (past month), with<sup>b</sup>: |  |
| Regular non-paying male partner | 935/2033 (46.0) |
| Paying male partners | 884/2375 (37.2) |
| Paid male partners | 310/885 (35.0) |
| Casual non-paying male partners | 483/1146 (42.1) |
| Any type of male partner | 1502/3328 (45.1) |

<sup>a</sup> Percentages may not add to 100 due to missing values.
<sup>b</sup> Among those who had that type of partner(s) and reported anal sex during the past month.

### Co-occurrence of adverse psychosocial exposures

Among the 4,607 HIV-negative transgender women, irrespective of other exposures, more than one-fourth (27.3%) reported having been exposed to physical violence and 22.3% to sexual violence. More than one-third (39.2%) had been exposed to either physical or sexual violence and 10.5% to both physical and sexual violence. Over one-third (33.9%) reported frequent alcohol use and 11.5% reported drug use. Table 3 reports the co-occurrence of physical violence, sexual violence, frequent alcohol use, and drug use. Overall, 16.5% (n=698) reported two co-occurring psychosocial exposures, ranging from 1.8% (n=76) co-occurrence of alcohol and drug use to 6.1% (n=256) alcohol use and physical violence. Three exposures were reported by 5.8% (n=243), and 2.7% (n=113) reported all four exposures. About 43% of participants reported no adverse psychosocial exposures.

**Table 3.** Adverse Psychosocial Exposures among HIV-negative Transgender Women (N = 4,607): Physical Violence, Sexual Violence, Frequent Alcohol Use, Drug Use, and their Co-occurrence.

| **Adverse psychosocial exposures and their co-occurrence** | **n (%)<sup>a</sup>** |
| --- | --- |
| Each exposure irrespective of other three exposures |  |
| Physical violence, past 12 months | 1257 (27.3) |
| Sexual violence, past 12 months | 1029 (22.3) |
| Drug use, past 12 months | 488 (11.5) |
| Frequent alcohol use, past week (>3 days/week) | 1563 (33.9) |
| Each exposure alone |  |
| Physical violence | 352 (8.3) |
| Sexual violence | 264 (6.2) |
| Drug use | 86 (2.0) |
| Frequent alcohol use | 655 (15.5) |
| Co-occurrence of exposures |  |
| Frequent alcohol use + Drug use | 76 (1.8) |
| Frequent alcohol use + Physical violence | 256 (6.1) |
| Frequent alcohol use + Sexual violence | 124 (2.9) |
| Physical violence + Drug use | 35 (0.8) |
| Sexual violence + Drug use | 62 (1.5) |
| Physical violence + Sexual violence | 145 (3.4) |
| Physical violence + Drug use + Frequent alcohol use | 40 (1.0) |
| Sexual violence + Drug use + Frequent alcohol use | 59 (1.4) |
| Physical violence + Sexual violence + Frequent alcohol use | 127 (3.0) |
| Physical violence + Sexual violence + Drug use | 17 (0.4) |
| Physical violence + Sexual violence + Frequent alcohol use + Drug use | 113 (2.7) |
| No adverse psychosocial exposures | 1814 (42.9) |

<sup>a</sup> Percentages may not add to 100 due to missing values

### Perpetrators of physical or sexual violence

Among those who experienced sexual violence (n=1,029), over one-third (35.9%; n=369) reported more than one type of perpetrator. Perpetrators were identified as strangers (41.1%), thugs (23.9%), clients (23.2%), and family members (21.8%). Among those who experienced physical violence (n=1,257), 32.5% (n=409) reported more than one perpetrator type. Perpetrators were identified as strangers (33.0%), police (24.5%), thugs (21.2%), family members (20.0%), and clients (17.3%).

### Synergistic interactions between adverse psychosocial exposures

Using logistic regression to model CAS, without product terms, physical violence [PV] had a statistically significant association with CAS (aOR=1.94, 95% CI: 1.47–2.56; p<.001), while frequent alcohol use [A] (aOR = 1.32, 95% CI: .98–1.77; p=.07), drug use [D] (aOR =.86, 95% CI: .60–1.22; p=.40) and sexual violence [SV] (aOR = .97, 95% CI: .73–1.29; p=.86) did not have statistically significant associations with CAS. Among the other covariates included in the multivariable logistic regression model of main effects without product terms, higher levels of education (aOR=.93, 95% CI: .90–.97; p<.001), being unmarried (aOR=.64, 95% CI: .45–.92; p=.016), and nirvan gender-affirmative surgical status (aOR=.68, 95% CI: .52–.88; p=.003) were associated with lower odds of CAS, and forced sex during adolescence with higher odds of CAS (aOR=1.35, 95% CI: 1.02–1.78; p=.04).

Out of six logistic regression models that included main effects and two-way product terms, the following product terms were statistically significant: PV x D (Model 1) and PV x SV (Model 5) (Table 4). In the additional five logistic regression models that included main effects and two- and three-way product terms, the following were statistically significant: PV x D (Model 7), PV x A x D (Models 8, 9, 10, and 11), D x PV x SV (Model 10) and A x PV x SV (Models 11 and 12). In the final logistic regression model that included all possible product terms, including a four-way product term, only A x PV x SV was statistically significant. Assessment of interactions on the additive scale using linear probability regression models showed similar results (Table 5).

**Table 4.** Effect of Adverse Psychosocial Exposures on Condomless Anal Sex with Male Partners among HIV-negative Transgender Women: Measures of Multiplicative Two-/Three-/Four-way Interactions between Physical Violence, Sexual Violence, Drug Use, and Frequent Alcohol Use (N = 4,607)

| Adverse psychosocial exposures and product terms | Effects on condomless anal sex |  |  |  |  |  |  |  |  |  |  |  |
| --- | --- | --- | --- | --- | --- | --- | --- | --- | --- | --- | --- | --- |
|  | Model 1: | Model 2: | Model 3: | Model 4: | Model 5: | Model 6: | Model 7: | Model 8: | Model 9: | Model 10: | Model 11: | Model 12: |
|  | Models with one two-way product term at a time |  |  |  |  |  | All two-way product terms<br>PV x D<br>PV x A<br>SV x D<br>SV x A<br>PV x SV<br>A x D | All two-way +<br>one three-way product term<br>(PV x A x D) | All two-way +<br>two three-way product terms<br>(PV x A x D)<br>(SV x A x D) | All two-way +<br>three three-way product terms<br>(PV x A x D)<br>(SV x A x D)<br>(D x PV x SV) | All two-way +<br>four three-way product terms<br>(PV x A x D)<br>(SV x A x D)<br>(D x PV x SV)<br>(A x PV x SV) | All two-way +<br>All three-way +<br>four-way product terms<br>(PV x A x D)<br>(SV x A x D)<br>(D x PV x SV)<br>(A x PV x SV)<br>(A x D x PV x SV) |
|  | PV x D | PV x A | SV x D | SV x A | PV x SV | A x D |  |  |  |  |  |  |
|  | Estimated semi-elasticity (95% CI), p value |  |  |  |  |  |  |  |  |  |  |  |
| PV x D | .48<br>(.27, .69)*** |  |  |  |  |  | .47<br>(.23, .71)*** | -.17<br>(-.83, .48) | -.14<br>(-.79, .52) | -.39<br>(-1.20, .42) | -.25<br>(-.99, .49) | -.11<br>(-.88, .65) |
| PV x A |  | .13<br>(-.16, .42) |  |  |  |  | -.03<br>(-.28, .31) | -.08<br>(-.43, .27) | -.09<br>(-.45, .26) | -.09<br>(-.44, .26) | -.38<br>(-.84, .08) | -.36<br>(-.83, .11) |
| SV x D |  |  | -.02<br>(-.39, .35) |  |  |  | -.12<br>(-.53, .29) | -.21<br>(-.64, .22) | -.02<br>(-.54, .58) | -.13<br>(-.77, .52) | -.12<br>(-.75, .52) | -.04<br>(-.69, .61) |
| SV x A |  |  |  | -.01<br>(-.33, .30) |  |  | -.10<br>(-.43, .24) | -.06<br>(-.39, .26) | -.01<br>(-.35, .33) | .01<br>(-.33, .34) | -.38<br>(-.91, .14) | -.36<br>(-.89, .18) |
| PV x SV |  |  |  |  | .29<br>(.04, .53)* |  | .23<br>(-.04, .49) | .21<br>(-.06, .48) | .22<br>(-.05, .48) | .13<br>(-.18, .44) | .31<br>(-.76, .14) | -.28<br>(-.75, .19) |
| A x D |  |  |  |  |  | .08<br>(-.27, .43) | -.06<br>(-.49, .36) | -.40<br>(-1.0, .19) | -.23<br>(-.90, .44) | -.17<br>(-.81, .47) | -.11<br>(-.73, .51) | -.05<br>(-.69, .59) |
| PV x A x D |  |  |  |  |  |  |  | .69<br>(.38, 1.0)*** | .69<br>(.38, 1.0)*** | .65<br>(.29, 1.0)*** | .61<br>(.23, .99)** | .52<br>(-.04, 1.08) |
| SV x A x D |  |  |  |  |  |  |  |  | -.39<br>(-1.32, .54) | -.65<br>(-1.72, .43) | -.53<br>(-1.56, .49) | -.74<br>(-2.09, .61) |
| D x PV x SV |  |  |  |  |  |  |  |  |  | .51<br>(.06, .96)* | .34<br>(-.24, .93) | .06<br>(-1.13, 1.25) |
| A x PV x SV |  |  |  |  |  |  |  |  |  |  | .66<br>(.35, .97)*** | .63<br>(.27, .99)*** |
| A x D x PV x SV |  |  |  |  |  |  |  |  |  |  |  | .40<br>(-.64, 1.46) |

PV, physical violence in the past year; SV, sexual violence in the past year; D, drug use in the past year; A: frequent alcohol use in the past week. All models adjusted for age, education, marital status, sexual identity, forced sex experience during adolescence, HIV risk perception, knowledge HIV transmission risk, social support, and HIV program exposure. The estimates of the main effects are not shown. Interpretation of semi-elasticity: a semi-elasticity of .48, for example in Model 1, is interpreted as a 48% relative change in the probability of condomless anal sex that is associated with the joint effect of physical violence and drug use, above and beyond their independent associations with the outcome. \* $p < .05$ , \*\* $p < .01$ , \*\*\* $p < .001$ .

**Table 5.**
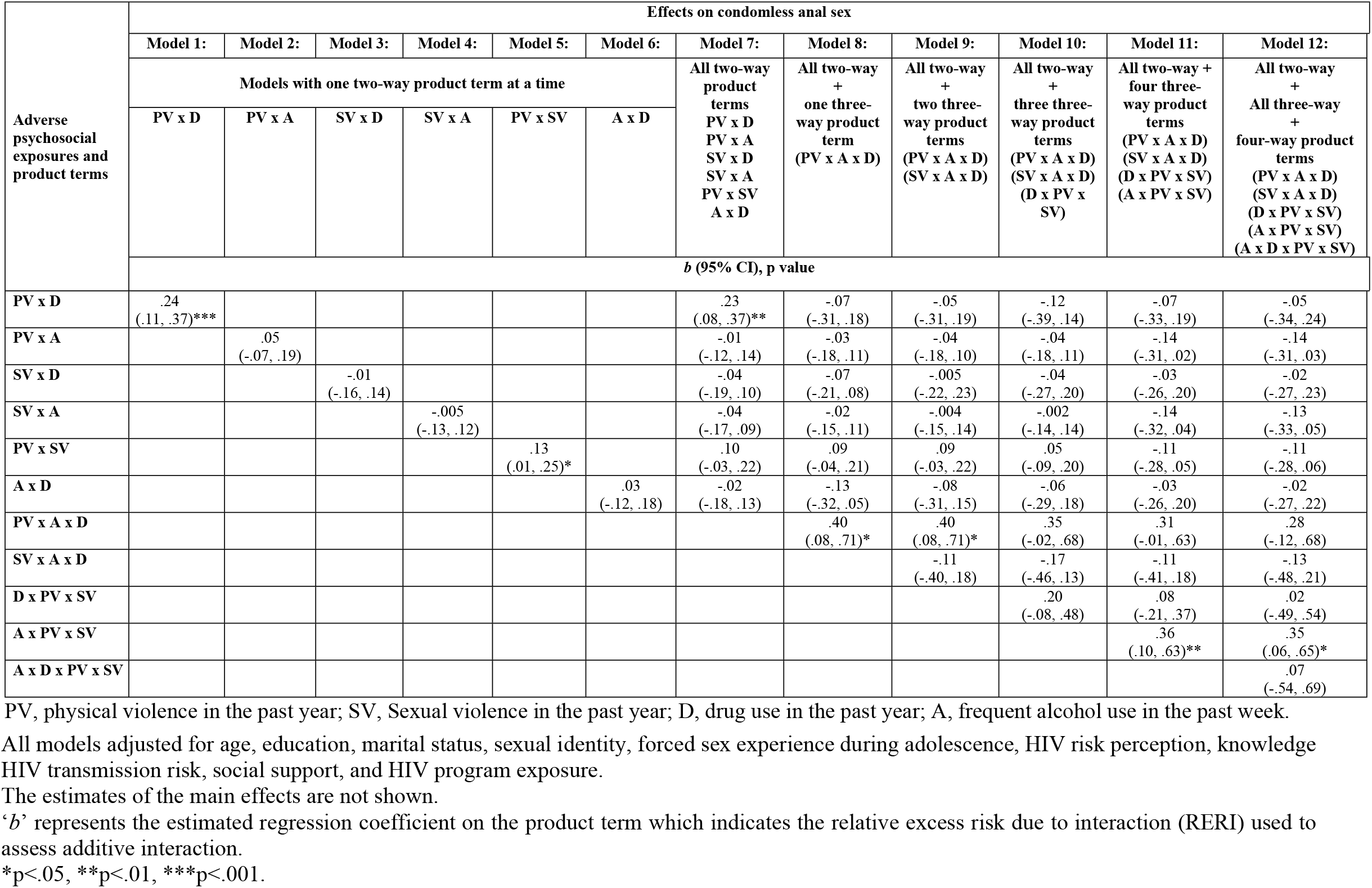
Effect of Adverse Psychosocial Exposures on Condomless Anal Sex with Male Partners among HIV-negative Transgender Women: Measures of Additive Two-/Three-/Four-way Interactions between Physical Violence, Sexual Violence, Drug Use, and Frequent Alcohol Use (n = 4,607)

### Sensitivity analysis for the model of synergistically interacting epidemics

The sensitivity analysis specifying drug use and alcohol use as continuous exposures, physical and sexual violence as a single continuous exposure (violence victimization), and CAS as a continuous outcome, showed a similar pattern of two- and three-way interactions (see S1 Table).

### Rival theoretical models: serially causal and mutually causal epidemics

In the model of serially causal epidemics, we estimated a statistically significant total indirect effect of sexual violence (estimate=.04, 95% CI: .003–07; p=.03) on CAS through drug use and alcohol use, but the indirect effect of physical violence was not statistically significant. In the model of mutually causal epidemics, we found that violence victimization had a statistically significant association with drug use (estimate=.18, 95% CI: .15–.22; p<.001) and drug use had a statistically significant association with violence victimization (estimate=1.97, 95% CI: 1.38–2.56; p<.001). The model fit (SRMR<.02) and the stability index (.60) were good (see S2 Table).

## Discussion

In this large, cross-sectional, probability-based analytical sample of HIV-negative transgender women in India, we tested a model of synergistically interacting epidemics, based on the joint effects of four psychosocial exposures—physical and sexual violence victimization, drug use, and alcohol use—on CAS. The high prevalence of these adverse psychosocial exposures, their frequent co-occurrence, and their synergistic interactions support the presence of synergistically interacting epidemics, or a syndemic. On both multiplicative and additive scales, we found evidence of two-way synergistic interactions between physical violence victimization and sexual violence victimization, and between physical violence victimization and drug use. We found evidence of three-way synergistic interactions between physical violence victimization, sexual violence, and alcohol use; and between physical violence victimization, drug use and alcohol use. Our findings suggest that a harmful social-structural environment for transgender women in India contributes to clustering of psychosocial health problems significantly associated with increased HIV risk, with ramifications for targeted HIV prevention initiatives.

The high levels of physical (27.3%) and sexual violence (22.3%) victimization faced by transgender women are corroborated by other studies from India [13,14,24]. A study from Karnataka reported past-year sexual violence prevalence of 18%, but did not disaggregate results among the combined sample of 543 men who have sex with men and transgender women [14]. In a study with 300 transgender women in four states, 84% reported having ever experienced physical and/or sexual violence [13]. Notably, in the present study, about one-third of those who experienced violence reported multiple perpetrators: family members/relatives, thugs, police, strangers, and sex work clients. Thus, not only do a substantial proportion of transgender women face violence; they experience violence across a myriad of settings in their social ecology—family, work, community settings, and in interactions with law enforcement officers, ostensibly positioned to offer protection from violence. A latent class analysis of patterns of violence victimization among transgender women in India (n=299) indicated that the type (verbal, physical, or sexual) of violence victimization differentially influenced HIV risk; those who experienced police-perpetrated sexual assault vs. police-perpetrated physical violence had relatively higher risk of CAS [24]. However, that study did not assess synergistic effects, such as those identified in the present study between physical and sexual violence, nor their interactions with alcohol and drug use.

Studies among gender and sexual minority populations in other contexts [25,26] indicate that the presence of multiple types and domains of violence victimization is likely to substantially increase psychological distress. The present study further demonstrates that multiple forms of violence victimization against transgender women, and in conjunction with drug use and alcohol use, contribute to synergistically increasing HIV risk. In India, physical violence from family members has been shown to be a manifestation of non-acceptance of gender identity, with violence reportedly perpetrated as a form of disciplining [27]. Physical and sexual violence from police and thugs in India have been identified as even more common among those transgender women who engage in sex work [28]. The multiple forms and domains of violence victimization can be seen as the manifestation of systemic stigma and discrimination undergirded by lack of acceptance of transgender people in the family and society; and this is exacerbated by the lack of legal protections for those who engage in sex work [29].

The associations identified using models of synergistically interacting epidemics or mediational analyses have been demonstrated among transgender women in other countries. For example, studies among transgender women in North and South America indicate that life stressors [30], discrimination [31,32] and trauma [33] can lead to alcohol and/or substance use, possibly as maladaptive coping strategies. A longitudinal study among transgender women in the U.S. showed a causal association between sexual violence and drug use [34]. In several other studies with transgender women, alcohol use [35] and drug use [36], including sexualized drug use [37], were associated with increased HIV risk. As identified among other populations, such as gay and bisexual men, being under the influence of alcohol or drugs may increase the likelihood of engaging in sex and the risk of CAS [38,39]. Thus, experiences of sexual violence can precipitate drug or alcohol use as a coping mechanism, and alcohol and drug use (especially before or during sex) in turn may increase the likelihood of being targeted or otherwise exacerbate sexual or physical violence from male partners [37], which increases the risk of engaging in CAS. Given evidence that internalized transprejudice and depression may stem in part from discrimination and violence victimization [40,41], future studies should explore the potential mediating roles of internalized transprejudice and depression in the association between physical and sexual violence victimization and discrimination, and HIV transmission risk behavior.

### Implications for practice and policy

Given that larger societal forces, including negative attitudes and systemic stigma and discrimination against transgender persons, and criminalization of sex work, contribute to violence victimization—as well as indirectly leading to alcohol and drug use through internalized stigma and depression [11,29,42]—addressing these social and structural factors is crucial. Currently, ‘crisis response’ teams established in NACO-supported targeted HIV interventions act only *after* incidents of violence are reported; and we are aware of no societal-level campaigns in India designed to prevent discrimination and violence against transgender women in relation to their intersecting marginalized identities (e.g., gender minority *and* sex worker and/or HIV-positive status). Important progress is evidenced in the passage of The Transgender Persons (Protection of Rights) Act, 2019 [43] and Rules, 2020 [44], in India, which explicitly states that transgender persons should not be discriminated against and prescribes punishment for those who discriminate or perpetrate violence. Nevertheless, disparities such as punishments for sexual violence against transgender women being less severe than that for cisgender women highlight ongoing challenges with protective policies and the devaluation of transgender women’s rights and lives. It is also vital to ensure the implementation of these policies on the ground.

Our findings further indicate that physical violence victimization independently, and together with sexual violence, drug use and alcohol use synergistically increase CAS. That is, in the presence of physical violence victimization, relatively higher levels of CAS were observed with one or more combinations of sexual violence victimization, drug use and/or alcohol use. Thus, addressing all four exposures through integrated, multicomponent interventions may substantially reduce HIV risk [45]; however, in the context of synergistic interactions, single-component interventions (e.g., post-care support for survivors of sexual violence or treatment of alcohol or drug dependence) may still exert an incremental impact on reducing HIV transmission risk behavior.

Partial support for the model of serially causal epidemics—i.e., sexual violence victimization leading to alcohol and drug use, and possible mutually causal associations between violence victimization and drug use—suggest the utility of multilevel interventions: at the individual level, screening for and managing alcohol and drug use; and at the societal level, strengthening stigma and violence reduction or elimination programs. Although alcohol use and drug use were not independently statistically significant predictors of CAS (main effects model), in the presence of sexual and/or physical violence victimization, alcohol use and/or drug use substantially increased CAS. Nongovernmental (NGOs) and community-based organizations (CBOs) that implement targeted HIV preventive interventions for transgender women should establish or strengthen referral systems with public hospitals offering treatment for alcohol and drug dependence, post-violence (physical or sexual) support services, and post-sexual exposure prophylaxis. To this end, training and monitoring healthcare providers to ensure nondiscriminatory and gender-affirmative care is crucial. Similarly, support is required from police in duly filing complaints of sexual or physical violence, given that transgender persons are not covered in current sexual assault/rape laws (Indian Penal Code, Section 354)— a pernicious lapse in legal policy. Further, in addition to HIV and condom education and condom distribution, NGOs and CBOs should educate transgender women about the negative effects of alcohol and drug use on sexual decision-making and negotiation, counselling and supporting them to reduce or avoid drug or alcohol use before or during sex.

### Limitations and strengths

In addition to its strengths, this study has several limitations. First, the cross-sectional data preclude causal interpretations. However, theoretical frameworks and compatible recall periods for the adverse psychosocial exposures (physical and sexual violence victimization, one year; drug use, one year), support the plausibility of the three models tested. Second, our analysis was restricted to co-occurrence and interactions between exposures at the individual level, while the concept of a syndemic is a population-level phenomenon; multilevel analyses at the population level should be undertaken in future research. Nevertheless, this is among the largest probability-based studies of syndemics among transgender women [46], and had a high response rate (81%). Our use of appropriate statistical analyses (interactions on both additive and multiplicative scales) to assess synergy is another strength, given several studies that purportedly assessed syndemics and HIV risk among transgender women did not use such analytic techniques [46], among very few that did [47]. In line with current practice, we dichotomized the adverse psychosocial exposures in additive and multiplicative interactions [18]; however, sensitivity analyses revealed that similar findings were obtained if we used different ways of specifying exposures, strengthening the evidence for synergy among exposures.

## Conclusion

This study found evidence for synergistic (additive and multiplicative) interactions between sexual and physical violence victimization, drug use, and alcohol use on HIV transmission risk behavior among transgender women. Our findings support the need to focus on preventing and eliminating violence victimization, as well as the importance of preventing or mitigating multiple co-occurring adverse psychosocial exposures, in order to reduce HIV risk among transgender women in India.

## Data Availability

The data underlying the results presented in the study are available from Dr. Shobini Rajan, National AIDS Control Organisation (NACO), India, who can be contacted at. Access will be granted to de-identified datasets on a case-by-case basis.

